# RT-qPCR detection of SARS-CoV-2 mutations S 69-70 del, S N501Y and N D3L associated with variants of concern in Canadian wastewater samples

**DOI:** 10.1101/2021.05.20.21257536

**Authors:** Shelley W. Peterson, Ravinder Lidder, Jade Daigle, Quinn Wonitowy, Audra Nagasawa, Michael R. Mulvey, Chand S. Mangat

## Abstract

SARS-CoV-2 variants of concern (VoC) have been increasingly detected in clinical surveillance in Canada and internationally. These VoC are associated with higher transmissibility rates and in some cases, increased mortality. In this work we present a national wastewater survey of the distribution of three SARS-CoV-2 mutations found in the B.1.1.7, B.1.351, and P.1 VoC, namely the S-gene 69-70 deletion, N501Y mutation, and N-gene D3L. RT-qPCR allelic discrimination assays were sufficiently sensitive and specific for detection and relative quantitation of SARS-CoV-2 variants in wastewater to allow for rapid population-level screening and surveillance. We tested 261 samples collected from 5 Canadian cities (Vancouver, Edmonton, Toronto, Montreal, and Halifax) and 6 communities in the Northwest Territories from February 16th to March 28th, 2021. VoC were not detected in the Territorial communities, suggesting the absence of VoC SARS-CoV-2 cases in those communities. Percentage of variant remained low throughout the study period in the majority of the sites tested, however the Toronto sites showed a marked increase from ~25% to ~75% over the study period.

The results of this study highlight the utility of population level molecular surveillance of SARS-CoV-2 VoC using wastewater. Wastewater monitoring for VoC can be a powerful tool in informing public health responses, including monitoring trends independent of clinical surveillance and providing early warning to communities.

## 1. Introduction

SARS-CoV-2, the causative agent of COVID-19, is a single-stranded RNA virus with the potential to undergo mutations that can lead to an increased viral transmission rate, increased virulence, or the ability to escape existing vaccines (Gorbalenya Alexander et al. (2020)). Since the fall of 2020, variants of concern (VoC) began to emerge which had higher infection rates and in some cases, increased mortality (Frampton et al. (2019)). These included the B.1.1.7 variant first described in the UK (Volz et al. (2021)), the B.1.351 variant first described in South Africa (Tegally et al. (2020)), and the P.1 variant first described in Brazil (Faria et al. (2021); Naveca et al. (2021)). The emergence and rapid dissemination of the B.1.1.7 variant has recently been reported in Toronto, Canada (Brown et al. (2021)).

Wastewater-based epidemiology (WBE) has emerged as a powerful tool to assist in the surveillance of SARS-CoV-2 (Hamouda et al. (2021)). The RT-qPCR assays developed for detection of COVID-19 clinical cases have now been adapted to detect and quantify SARS-CoV-2 RNA in wastewater. RT-qPCR has been demonstrated to detect the presence of the virus in wastewater prior to identification of clinical cases in some regions, which highlights the usefulness of the assay as a cost-effective early warning system for some communities (Farkas et al. (2020)). In addition, quantification of the virus over time can be used to monitor trends, aiding in assessing the effectiveness of public health interventions designed to reduce viral transmission (Farkas et al. (2020)).

There are a number of methods that can be employed for VoC detection in wastewater including next generation sequencing and RT-qPCR (Fontenele et al. (2021)). Although next generation sequencing has the potential to identify both extant and emerging variants in wastewater, the process has longer turn around times, is costly, and requires complex infrastructure and bioinformatics expertise (Yaniv et al. (2021)). Alternatively, building on the success of RT-qPCR-based assays for SARS-CoV-2 detection in wastewater, RT-qPCR-based assays have been developed to identify mutations associated with emerging VoC including Sdel69-70 (B.1.1.7), and Sdel241 (B.1.351) (Yaniv et al. (2021)). In addition, a primer extension assay has been described to identify the ND3L (B.1.1.7) variant in wastewater (Graber et al. (2021)). Although these assays are based on lineage specific diagnostic mutations, confirmation by sequencing is required to definitively identify true VoC in a population. The benefits of rapid identification of SARS-CoV-2 VoC in wastewater are potential early public health responses such as increased sequencing of clinical isolates, enhanced surveillance, and enhanced public health measures. In this report we validate multiple RT-qPCR assays directed to specific alleles associated with the SARS-CoV-2 B.1.1.7, B.1.351, and P.1 variants in wastewater. The multiple targets described herein will provide a specific multi-locus assay for the early detection of variants in wastewater samples potentially identifying the variant prior to clinical findings similar to what has previously been described for the wild-type (WT) SARS-CoV-2 in wastewater (reviewed in(Hamouda et al. (2021))). We applied this assay to longitudinal wastewater samples collected from 22 sampling locations drawn from 11 Canadian cities and towns, both remote and urban, to develop a national picture of the emergence of B.1.1.7, B.1.351, and P.1 variants in wastewater relative to wild-type. The approach described here could be utilized by public health officials to monitor changes in prevalence of the virus and monitor interventions to reduce the number of infections in a health region, and inform areas refractory to clinical surveillance.

## 2. Materials and Methods

### 2.1 Wastewater collection

Using the infrastructure provided by Statistics Canada’s Canadian Wastewater Survey (CWS), wastewater was collected from fifteen urban (Vancouver, Edmonton, Toronto, Montreal and Halifax) wastewater treatment plants (WWTP). Wastewater was also collected from 7 lift stations from remote communities in the Northwest Territories that are not part of the CWS. In total, the wastewater examined in this study represents over 23% of the Canadian population. A 24 hr composite sample was collected three times per week at each treatment facility and shipped to the National Microbiology Laboratory at 4 °C. The samples were stored at 4 °C for up to 24 hrs until processed.

### 2.2 Sample processing and RNA extraction

A 300 mL 24-hour composite sample of primary post-grit influent or raw wastewater was mixed by inversion and then a 30 mL aliquot was drawn and centrifuged at 4,000 x g for 20 minutes at 4 °C. The supernatant was removed using a pipette, leaving approximately 500 µL of supernatant with the pellet. The pellet was resuspended and transferred to a 2 mL tube containing 200 µL 0.5 mm zirconia-silicate beads (Biospec, Bartlesville, OK) and 700 µL Qiagen Buffer RLT (Qiagen, Germantown, MD) containing 1% 2-mercaptoethanol. The mixture was processed in a Bead Mill 24 Homogenizer (Fisher Scientific, Ottawa, ON) at 6 m/s for four 30 second cycles. The pelleted debris was removed by centrifugation at 12,000 x g for 3 minutes, and up to 1 mL lysate was transferred to a new 2 mL processing plate containing 2 µL of 10 mg/µL carrier RNA (Sigma-Aldrich, Oakville, ON) in each well. RNA was extracted using the Magna Pure 96 DNA and Viral NA Large Volume Kit (Roche Diagnostics, Laval, QC) using the Plasma External Lysis 4.0 protocol as per manufacturer instructions.

### 2.3 Assay Design

Assays were developed to detect B.1.1.7, B.1.351 and P.1 based on the high profile and global distribution of these lineages and their association with high SARS-CoV-2 transmissibility (Leung et al. (2021); Ramanathan et al. (2021)). The spike protein N501Y mutation consisting of an A->T at position 23063 alters one of the key contact residues within the receptor binding domain, leading to increased binding affinity to ACE2 in humans (Ramanathan et al. (2021); Rynkiewicz et al. (2021)). This mutation is found in B.1.1.7, B.1.351, and P.1; and is believed to have arisen independently in both lineages (Martin et al. (2021)). Another clinically relevant mutation, a deletion of amino acid residues 69-70 in the spike protein (position 21765-21770), has led to spike gene target failure in some clinical diagnostic assays in addition to increased infectivity in B.1.1.7 (Bal et al. (2020); Kemp et al. (2021)). The N D3L mutation, consisting of a GAT->CTA mutation at position 28280 in B.1.1.7 was chosen due to close proximity to the N1 RT-qPCR assay currently in use for detection of SARS-CoV-2 in wastewater. To further support the specificity of these SNPs, an investigation of 20,000 Canadian SARS-CoV-2 sequences collected prior to the detection of B.1.1.7, B.1.351, and P.1 in Canada, occurrences of S N501Y, S 69/70 del, and N D3L were found to be 0.019%, 0.028%, and 0.014% respectively, thus making them effective targets for variant detection (G. Van Domselaar, personal communication).

Due to the high complexity and variability of the wastewater matrix, this assay was designed to detect both WT and variant (V) sequences for each allele, allowing for relative quantitation of variant to WT SARS-CoV-2 RNA within the sample.

### 2.4 RT-qPCR Allelic Discrimination

The wild-type SARS-CoV-2 (NC_045512.1) sequence, along with variant sequences B.1.1.7 (EPI_ISL_742238), B.1.351 (EPI_ISL_736935), and P.1 (EPI_ISL_1445274) were obtained from GISAID (www.gisaid.org) and used for primer and probe design. Two gene targets were chosen from the S gene: a deletion at 69-70 (Sdel) associated with B.1.1.7, and the N501Y mutation (SN501Y) associated with B.1.1.7, B.1.351, and P.1, along with one N gene mutation (D3L) associated with B.1.1.7. Oligonucleotide primers and probes were chosen for each target region using Primer Express Software v3.0 (ThermoFisher Scientific, Waltham, MA) and Primer3 v4.1.0 (https://bioinfo.ut.ee/primer3/). The primers and probes for each of the target mutations are shown in Table 1.

**Table 1:**
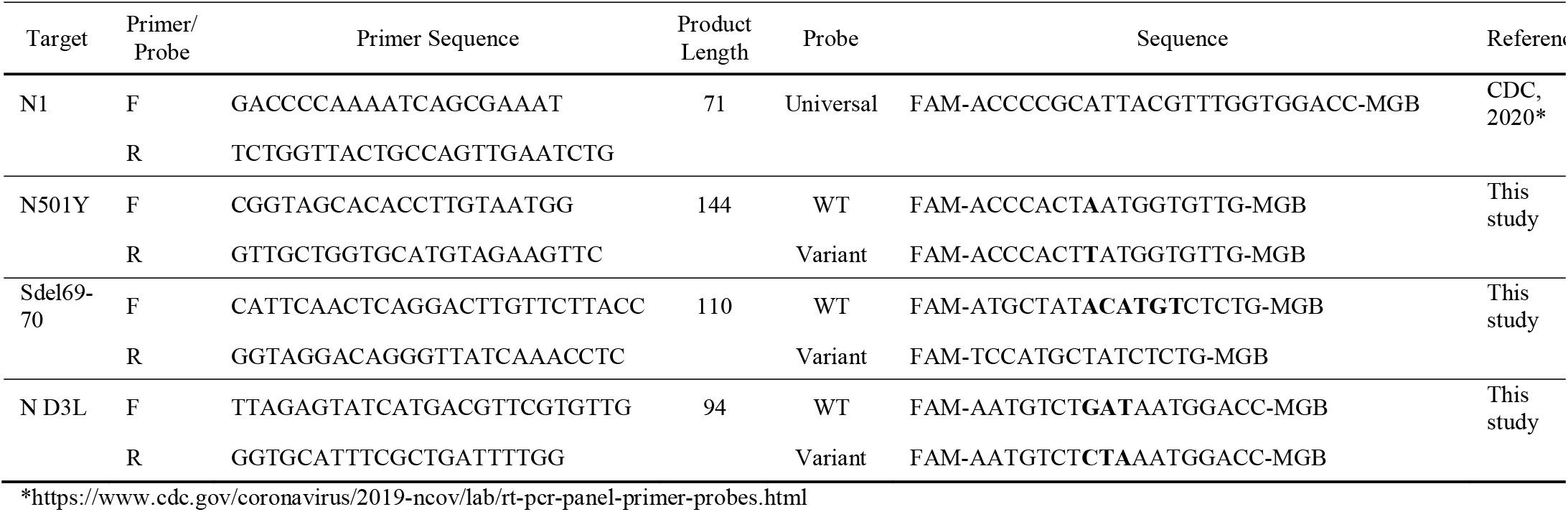
SARS-CoV-2 RT-qPCR Allelic Discrimination Primers and Probes

RT-qPCR was performed in a reaction volume of 20 µL, consisting of 5 µL TaqPath 1-Step RT-qPCR Master Mix, CG (ThermoFisher Scientific), 500 nM (ND3L), 400 nM (Sdel), 250 nM (N501Y) final concentration of each primer (ThermoFisher Scientific), 500 nM (D3L, Sdel), 250 nM (N501Y) of each probe (ThermoFisher Scientific), 4 µL of template RNA and Invitrogen nuclease free H_2_O (ThermoFisher Scientific). PCR amplification and detection of amplification products were performed on a QuantStudio 5 Real-time PCR instrument (ThermoFisher Scientific). Thermal cycling conditions were as follows: initial UNG incubation at 25 °C for 2 min, reverse-transcriptase incubation at 50 °C for 15 min, polymerase activation at 95 °C for 2 min; 40 cycles of 95 °C for 5 s and 60 °C for 30 s; and a final elongation step of 60 °C for 30 s. Results were considered positive if they had a Ct value <40 (Bustin et al. (2009)) and a visible amplification curve on the multicomponent plot using the QuantStudio 5 Design & Analysis Software (Thermo Fisher Scientific). Each real-time PCR was performed in duplicate or triplicate as indicated with the appropriate non-template controls and positive controls.

### 2.5 Limit of Detection

Limits of detection (LOD) were determined in triplicate using serial dilutions from 10^4^ to 10^0^ cp/µl of dsDNA oligonucleotide gene fragments (IDT, Coralville IA) made up of the gene region flanking the variant region for either the WT or variant sequence (Table S2). Six different DNA oligonucleotides were used, with individual WT and variant genotypes for each locus tested.

## 3. Results

### 3.1 Assay Limit of Detection, Stability, Specificity and Variance

The limits of detection were assessed at 100% test positivity in triplicate and determined to be 1 copy (cp)/µL for all targets when measured as a pure specimen without interfering alleles. The amplification efficiencies of each primer and probe set were between 90% and 102% (Table 2). As relative amounts of WT and V template within wastewater samples may vary considerably, standard curves were also created for each assay in the presence of 100, 500, and 1000 cp/µL of the alternate allele to assess their stability (Figure 1). There was no difference in LOD for the SN501Y assays in the presence of the alternate allele, however the Sdel and ND3L assays had an LOD of 10 cp/µL in the presence of 500 and 1000 cp/µL of the alternate allele, with the ND3L WT assay LOD increasing to 10 cp/µL in the presence of 100 cp/µL alternate allele as well. In addition, all assays except SN501Y WT were inhibited at a 10:1000 cp/µL ratio of target:alternate allele, demonstrating a ~2-4 Ct increase compared with Cts in the absence of the alternate allele. The standard deviation of the alternate allele in this experiment was used to assess assay variance in the presence of a range of concentrations of the alternate template. Standard deviations for each assay were averaged across the three concentrations and were as follows: 0.09 Ct (ND3L WT), 0.12 Ct (ND3L V), 0.15 Ct (Sdel WT), 0.09 Ct (Sdel V), 0.18 Ct (SN501Y WT) and 0.17 Ct (SN501Y V).

**Table 2:** Standard curve summaries for RT-qPCR SARS-CoV-2 variant detection and the CDC N1 assay. PCR efficiency and y-intercept cycle threshold (Ct) values were calculated for each of the primer-probe sets against tenfold dilutions of the corresponding DNA oligonucleotides. N1 was tested with both the WT and B.1.1.7 oligonucleotides.

|  | **Sdel WT** | **Sdel V** | **SN501Y**<br>**WT** | **SN501Y**<br>**V** | **ND3L**<br>**WT** | **N1 (WT**<br>**template)** | **ND3L V** | **N1 (V**<br>**template)** |
| --- | --- | --- | --- | --- | --- | --- | --- | --- |
| **Slope** | -3.43 | -3.36 | -3.56 | -3.39 | -3.47 | -3.57 | -3.28 | -3.34 |
| **Intercept** | 38.60 | 37.88 | 39.70 | 38.76 | 38.26 | 36.65 | 37.48 | 35.75 |
| **RSQ** | 1.000 | 0.996 | 0.994 | 0.999 | 0.997 | 1.000 | 0.999 | 0.998 |
| **Efficiency** | 95.54 | 98.40 | 90.89 | 97.16 | 94.27 | 90.48 | 101.67 | 99.28 |

**Figure 1:**
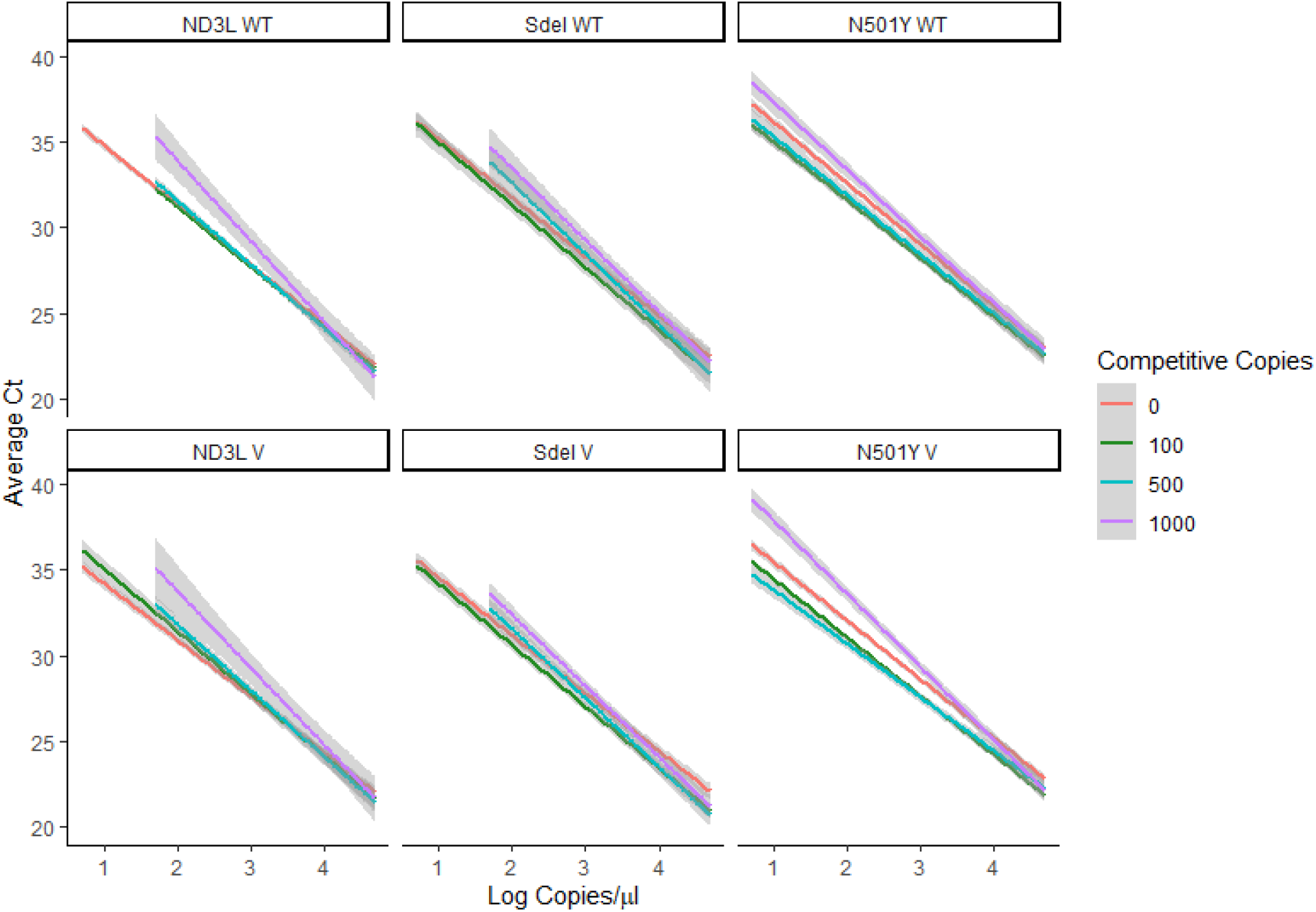
Standard curves for RT-qPCR SARS-CoV-2 variant assays against tenfold dilutions of DNA oligonucleotide controls with either the wild-type (WT) or B.1.1.7 (V) genotype in the presence of 100, 500, and 100 copies/µL of the alternate genotype for each locus.

To test analytical specificity, all variant targets were tested in triplicate with 100 cp/µL of the corresponding WT oligonucleotides and all WT targets were tested with 100 cp/µL of the corresponding variant oligonucleotides. No cross-reactivity was observed for any of the targets.

### 3.2 Assay comparison with CDC N1 in wastewater

When comparing qualitative detection of SARS-CoV-2 in wastewater with N1 as the gold standard, the sensitivities and specificities were 85.7% and 89.8% for ND3L, 96.1% and 93.4% for Sdel, and 96.1% and 95.2% for SN501Y, respectively. A total of 261 samples from fifteen urban WWTPs and 7 lift stations from remote communities across Canada were sampled from February 16 to March 28, 2021. Sites were tested 2-18 times with a median of 13 samples per site (Table 3). All 261 samples were tested for SARS-CoV-2 variants using the Sdel and SN501Y assays, and a subset of 140 samples were tested with both the S-gene assays and the ND3L assay (Table S1). Of these, 83/140 (59.3%), 176/261 (67.4%), and 175/261 (66.1%) tested positive for SARS-CoV-2 using ND3L, Sdel, and SN501Y WT or VoC targets, respectively, compared with 91/140 (65.0%) and 178/261 (68.2%) detected using the CDC N1 target as the gold standard.

**Table 3:** Location and number of samples tested from 5 municipalities and 7 lift stations in the Northwest Territories in Canada

| Site Code | Region | Sampling Date Range | No. of Samples |
| --- | --- | --- | --- |
| E1 | Edmonton | 2021-03-02 to 2021-03-21 | 9 |
| H1 | Halifax | 2021-02-22 to 2021-03-26 | 15 |
| H2 | Halifax | 2021-03-01 to 2021-03-26 | 12 |
| H3 | Halifax | 2021-02-22 to 2021-03-26 | 15 |
| M1 | Montreal | 2021-02-16 to 2021-03-28 | 18 |
| M2 | Montreal | 2021-02-16 to 2021-03-28 | 18 |
| T1 | Toronto | 2021-02-16 to 2021-03-23 | 13 |
| T2 | Toronto | 2021-02-16 to 2021-03-23 | 13 |
| T3 | Toronto | 2021-02-16 to 2021-03-23 | 13 |
| T4 | Toronto | 2021-02-16 to 2021-03-23 | 10 |
| V1 | Vancouver | 2021-02-16 to 2021-03-18 | 14 |
| V2 | Vancouver | 2021-02-16 to 2021-03-18 | 14 |
| V3 | Vancouver | 2021-02-16 to 2021-03-18 | 14 |
| V4 | Vancouver | 2021-02-16 to 2021-03-18 | 11 |
| V5 | Vancouver | 2021-02-16 to 2021-03-18 | 13 |
| N1 | Northwest Territories (NWT) | 2021-02-24 to 2021-03-24 | 9 |
| N2 | Northwest Territories (NWT) | 2021-03-17 to 2021-03-24 | 2 |
| N3 | Northwest Territories (NWT) | 2021-02-24 to 2021-03-26 | 11 |
| N4 | Northwest Territories (NWT) | 2021-03-08 to 2021-03-25 | 5 |
| N5 | Northwest Territories (NWT) | 2021-03-01 to 2021-03-25 | 7 |
| N6 | Northwest Territories (NWT) | 2021-02-26 to 2021-03-26 | 13 |
| N7 | Northwest Territories (NWT) | 2021-03-01 to 2021-03-26 | 12 |

Results for all three loci were in agreement for 123/140 (87.8%, 76 positive, 45 negative) samples tested with all 6 assays, and both S gene loci were in agreement for 254/261 (97.3%, 172 positive, 82 negative) samples. Four samples (3 from the Halifax region, 1 from Northwest Territories (NWT)) were negative with all variant assays tested and positive for N1, while three samples from Halifax had SARS-CoV-2 detection with all variant assays tested and no N1 detection. The concentration of SARS-CoV-2 detected in these Halifax and NWT regions was low (<10 cp/mL) for all of these samples. Ct values for these assays were consistently 2-3 Ct higher than N1 when testing wastewater, rendering them more suitable for relative quantitation of variant to WT alleles than absolute quantification of SARS-CoV-2 in wastewater.

### 3.3 Detection of SARS-CoV-2 Variants in Wastewater

SARS-CoV-2 RNA was detected by at least one of these six assays in all 15 urban sites and 2/7 remote lift stations (N3 – 2021-03-05, N7 - 2021-02-26). Variant allele percentages detected in each site are shown in Figure 2.

**Figure 2:**
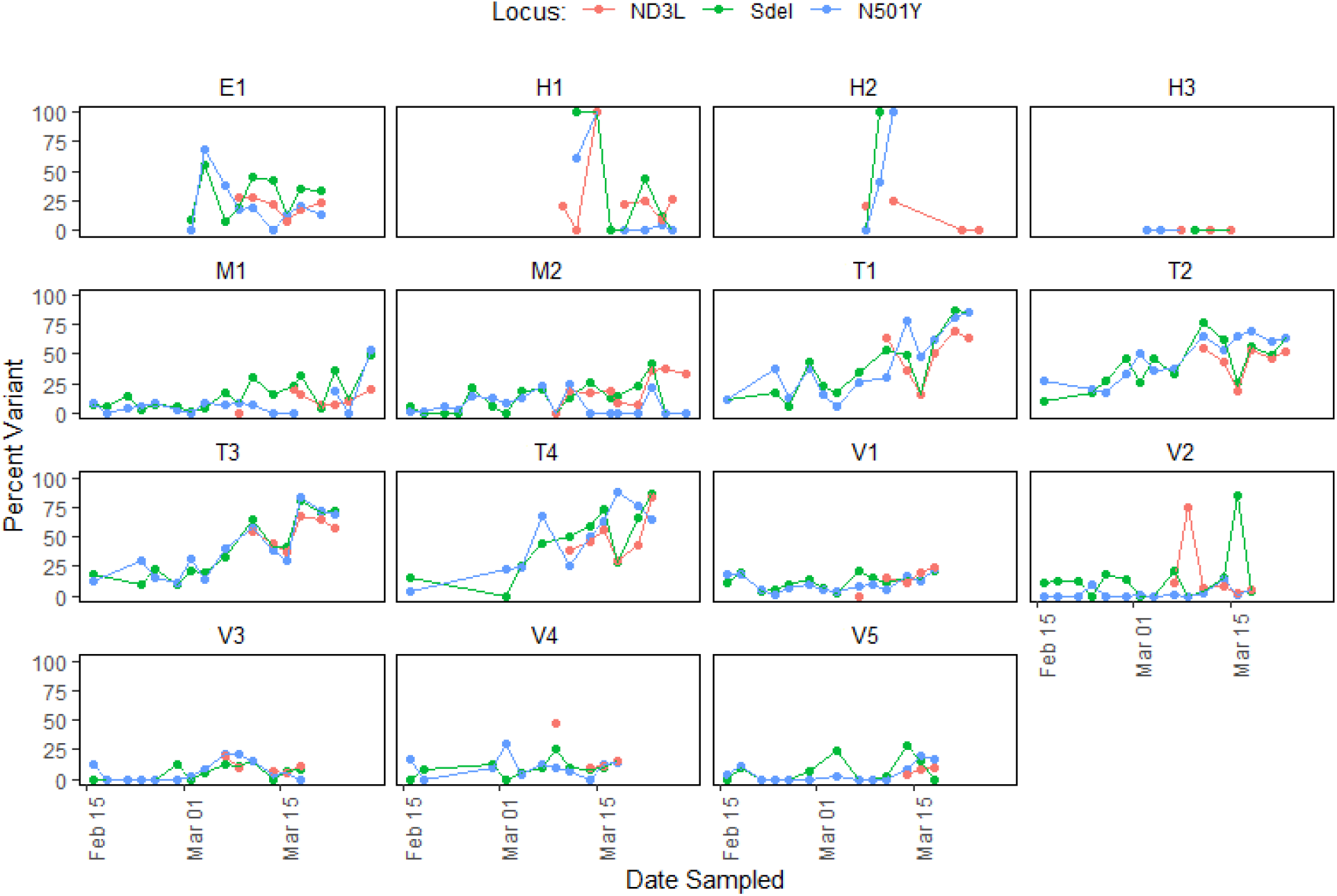
Percentage of SARS-CoV-2 variant alleles detected using RT-qPCR in wastewater across Canada from February 16 to March 28, 2021.

Of the three Halifax sites (H1-H3), no variant was detected during any time point in H3, and S-gene variants were detected between 2021-03-08 and 2021-03-12 in H2. All three variant mutations were detected in varying amounts in H1 from 2021-03-10 to 2021-03-26 with the exception of 2021-03-17. In both H1 and H2, variant percentages were inconsistent, varying widely (0-100% and 6.9-70.1%, respectively) possibly reflecting the low incidence at these sites, leading to less consistent detection of each allele.

The Edmonton site (E1) showed a large spike in B.1.1.7 variant detection (average 62.3%) on 2021-03-04, followed by consistent detection ~20-30%. Both Montreal sites (M1, M2) showed consistent B.1.1.7 variant detection, with the exception of one time point (2021-03-09) for M2 in which no variant was detected. In M1, variant detection was consistently low from 2021-02-26 to 2021-03-07, at which point there was an increasing trend with an average of 41.1% B.1.1.7 variant detection at 2021-03-28. In M2, percentages were more variable, ranging from average 1% to 33.4% throughout the study period.

For the five Vancouver sites (V1-V5), the relative percentage of variant showed no apparent increasing or decreasing trend. V1 and V4 had consistent B.1.1.7 variant detection across all time points with average variant percentages ranging from 3.1-22.9% (V1) and 3.8-27.4% (V4). N501Y was present at 12.5% at the V3 site on 2021-02-16, followed by two weeks without detection. Beginning on 2021-02-28, V3 showed consistent detection of at least one variant target with detection reaching >20% 2021-03-07 to 09 and then decreasing to below 10% for the remainder of the study period. B.1.1.7 variant targets were detected in V5 (average 1.5-14.1%) in the second and fourth weeks of February and the third week of March, with periods of no detection in between. The V2 site showed little agreement between the three alleles and a few larger spikes in percentage (75-85%) of the ND3L and Sdel targets.

All sites in the Toronto region (T1-T4) showed consistent detection across all time points along with upward trends from averages of 10.9%, 19.1%, 15.4%, and 9.1%, respectively at the first sampling time (2021-02-16) to 77.9%, 60%, 66.7%, and 78.3% at the last time point (2021-03-23). These data show that while the proportion of variant genotype to WT is rapidly increasing in the Toronto area, the variant allele had not yet fully replaced the wild-type during the study period in any of the sites tested from metropolitan areas with consistent SARS-CoV-2 present. In addition, there was no appreciable difference between percentage of N501Y signal detected compared with Sdel or ND3L in any sites, indicating that B.1.1.7 was the predominant variant in Canada throughout the study period.

No variants were detected in any of the NWT lift stations, corresponding with known epidemiological data in these regions (Government of Northwest Territories (2021)).

## 4. Discussion

The present study describes the development of three RT-qPCR assays to detect common SARS CoV-2 genomic alterations associated with the B.1.1.7, B.1.351, and P.1 VoC. These RT-qPCR assays provide sufficiently high sensitivity and specificity for detection and relative quantitation of the mutations in wastewater. These assays consist of six individual one-step RT-qPCR reactions that can easily be integrated into current wastewater surveillance protocols allowing for rapid and cost-effective detection and relative quantitation of mutations associated with variants of concern to aid in SARS-CoV-2 surveillance. In addition, this assay format is readily adaptable to additional emerging variants of concern by incorporating additional primers and probes as the need arises. Wastewater surveillance has been a highly valuable tool in for monitoring the spread of COVID-19 on a population level and has been instrumental in providing early warnings in care facilities and remote communities leading to public health action ((Aziz (2021); Burke and Romualdo (2020); CBC News (2020))). This study is the first temporal analysis investigating the relative proportion of mutations associated with VoC in wastewater across Canada.

Variant signals in wastewater were detected in all five major Canadian cities tested. In Toronto, the Canadian epicentre of B.1.1.7, the WT allele was predominant until mid-March, when the variant allele became more prevalent, representing up to 75% of signal detected. In regions with low prevalence, such as Halifax, where the concentration of SARS-CoV-2 RNA in the wastewater approaches the LOD of the assays, less reliable detection can lead to lower reliability in relative quantitation. However, in the Territorial sites no variants were detected, corresponding with known case data in those remote locations (Government of Northwest Territories). The absence of false negatives in these regions increases our confidence in the specificity of these assays.

There are many limitations to detection of SARS-CoV-2 variants in wastewater. Wastewater is a highly complex and variable matrix containing numerous potential inhibitors and organisms in addition to multiple SARS-CoV-2 variants increasing the necessity for highly specific assays. Furthermore, SARS-CoV-2 viral RNA is present in low concentrations in wastewater, necessitating the development of assays with low limits of detection. Presence of high concentrations of the alternate allele was shown to increase LODs of these assays, however detectable ratios as high as 1:500 are highly unlikely due to the low concentration of SARS-CoV-2 in Canadian wastewater (95% <250 cp/mL N1). Of the three loci for which assays were designed, the SN501Y assay was the most robust, with no loss of sensitivity in the presence of competing template and the highest sensitivities and specificities. In addition, the SN501Y mutation is present within multiple SARS-CoV-2 lineages (B.1.1.7, B.1.351, P.1. and others). In a region with limited resources, the SN501Y assay alone would be effective for detecting any of these variants. However, consistent with other SARS-CoV-2 variant detection assays (Graber et al. (2021); Yaniv et al. (2021)), these assays cannot replace the use of the CDC N1 assay for quantitative detection of SARS-CoV-2 due to ~2-3 Ct later detection. Finally, while lineage specific strategies for RT-qPCR based detection of VoC in wastewater can be informative to public health, the rapid emergence of multiple lineages necessitates directing assays toward genetic alternations that are both of medical importance and indicative of a wide-range of VoC. While detection of ND3L, Sdel or SN501Y are highly indicative of the presence of B.1.1.7, B.1.351, or P.1 they are not determinative, as these variants contain multiple co-occurring mutations that are not detected by this assay. However, a recent study detecting Sdel in wastewater from Paris found that the relative quantity of the mutation in wastewater was comparable to the estimated proportion of B.1.1.7 in positive patients (Wurtzer et al. (2021)). The relative proportion of B.1.1.7 to WT in wastewater can be estimated using the Sdel and ND3L assays, while N501Y is present in B.1.1.7, B.1.351, and P.1. Therefore, a high percentage of SN501Y detected with a lower percentage of ND3L and Sdel may indicate presence of B.1.351 or P.1 within a community. While whole genome sequencing is necessary for confirmation of the presence of variants, as well as discovery of new variants of concern, RT-qPCR provides a rapid cost-effective method of screening for variants at the population level. The results of this study highlight the utility of a molecular method of detection of mutations associated with the B.1.1.7, B.1.351, and P.1 SARS-CoV-2 variants in wastewater for population-level surveillance. The low LOD, low variation between assay replicates, and high specificity are proof-of-principle that this assay is an effective method of detecting the presence of variant and wild-type SARS-CoV-2 genetic material. The assays were used to test wastewater samples from urban and rural locations across Canada and determine the relative proportion of variant to WT viral RNA over a 7-week period. Variants were detected in five Canadian cities, with a high proportion of variant detected in the Toronto region. These tests may be beneficial for areas where these specific VoC have not been identified thus providing a cost-effective early warning system for communities. The data could also be used to aid in the identification of samples for more costly and time consuming metagenomic sequence analysis.

## 5. Conclusion

- Wastewater screening is a powerful tool for population-level detection of SARS-CoV-2 variants and can be used to inform public health responses by providing early warning to communities.
- While next generation sequencing is required to confirm presence of variants in wastewater, RT-qPCR is quicker and more cost-effective, and therefore, advantageous for wastewater screening.
- We developed a molecular method of detection of mutations associated with the B.1.1.7, B.1.351, and P.1 SARS-CoV-2 variants in wastewater for population-level surveillance, and applied this method to investigate the relative proportion of mutations associated with VoC in wastewater across Canada.
- Incidence of B.1.1.7 increased appreciably in Toronto between February and March 2021.

## Data Availability

Data is available as supplemental material

## Acknowledgments

We would like to thank Stacie Langner, Umar Mohammed, Graham Cox, Codey Dueck, Dave Spreitzer, and Norman Barairo for their technical assistance and support. Gary Van Domselaar and the CanCoGen team for providing genomic information on circulating SARS-CoV-2 in Canada. The Government of the Northwest Territories departments of Municipal and Community Affairs (Justin Hazenberg) and Environment and Natural Resources – Taiga Environmental Laboratory (Diep Duong) for assistance with wastewater sampling. The communities of the Northwest Territories for providing wastewater for analysis. Dr. Heather Hannah & Dr. Kami Kandola from the Department of Health and Social Services – Government of the Northwest Territories.

## Funding

This work was supported by internal funds from the Public Health Agency of Canada.

## Declaration of Competing Interests

None to declare.

## Notes

### Competing Interest Statement

The authors have declared no competing interest.

### Clinical Trial

Not Applicable

